# ODIASP: An Open User-Friendly Software for Automated SMI Determination—Application to an Inpatient Population

**DOI:** 10.1101/2024.10.25.24316094

**Authors:** Katia Charrière, Antoine Ragusa, Béatrice Genoux, Antoine Vilotitch, Svetlana Artemova, Charlène Dumont, Paul-Antoine Beaudoin, Pierre-Ephren Madiot, Gilbert R. Ferretti, Ivan Bricault, Eric Fontaine, Jean-Luc Bosson, Alexandre Moreau-Gaudry, Joris Giai, Cécile Bétry

**Author notes:** <u>Corresponding author</u> Prof. Cécile Bétry, MD-PhD, Université Grenoble Alpes, CHU Grenoble Alpes (CHUGA), Service d’Endocrinologie Diabétologie Nutrition, Boulevard de la Chantourne, 38700 La Tronche, France.

## Abstract

**Background:** The diagnosis of malnutrition has evolved with the GLIM recommendations, which advocate for integrating phenotypic criteria, including muscle mass measurement. The GLIM framework specifically suggests using skeletal muscle index (SMI) assessed via CT scan at the third lumbar level (L3) as a first-line approach. However, manual segmentation of muscle from CT images is often time-consuming and infrequently performed in clinical practice. This study aims to develop and validate an open-access, user-friendly software tool called ODIASP for automated SMI determination.

**Methods:** Data were retrospectively collected from a clinical data warehouse at Grenoble Alpes University Hospital, including epidemiological and imaging data from CT scans. All consecutive adult patients admitted in 2018 to our tertiary center who underwent at least one CT scan capturing images at the L3 vertebral level and had a recorded height were included. The ODIASP tool combines two algorithms to automatically perform L3 slice selection and skeletal muscle segmentation, ensuring a seamless process. Agreement between cross-sectional muscle area (CSMA) values obtained via ODIASP and reference methodology was evaluated using the intraclass correlation coefficient (ICC). The prevalence of reduced SMI was also assessed.

**Results:** SMI values were available for 2,503 participants, 53.3% male, with a median age of 66 years [51-78] and a median BMI of 24.8 kg/m^2^ [21.7-28.7]. There was substantial agreement between the reference method and ODIASP (ICC: 0.971; 95% CI: 0.825 to 0.989) in a validation subset of 674 CT scans. After correcting for systematic errors (a 5.8 cm^2^ [5.4-6.3] overestimation of the CSMA), the agreement improved to 0.984 (95% CI: 0.982 to 0.986), indicating excellent agreement. The prevalence of reduced SMI was estimated at 9.1% overall (11.0% in men and 6.6% in women). To facilitate usage, the ODIASP software is encapsulated in a user-friendly interface.

**Conclusions:** This study demonstrates that ODIASP is a reliable tool for automated muscle segmentation at the L3 vertebra level from CT scans. The integration of validated AI algorithms into a user-friendly platform enhances the ability to assess SMI in diverse patient cohorts, ultimately contributing to improved patient outcomes through more accurate assessments of malnutrition and sarcopenia.

## Introduction

Since the release of the GLIM recommendations, malnutrition diagnosis has been based on both phenotypic and etiological criteria ^1^. In current clinical practice, phenotypic criteria for malnutrition primarily involve mainly Body Mass Index (BMI) and weight loss assessments ^2,3^. However, a large proportion of patients are not weighed during hospitalization ^4^. The GLIM framework recommends that muscle mass assessments should be performed as a first-line approach to detect muscle mass reduction using one of three methods: dual-energy x-ray absorptiometry (DEXA), computerized tomography (CT) scan, or bioelectrical impedance analysis (BIA). For CT, the cross-sectional muscle area (CSMA) at the third lumbar vertebra (L3) is recommended to calculate the skeletal muscle index (SMI). However, manual segmentation of muscle from CT images is time-consuming^3^.

AI algorithms have the potential to automate SMI determination, potentially extending the use of CT scan for malnutrition diagnosis in clinical practice and across large volumes of CT scans ^5^. Although research in AI-driven CT scan analysis is expanding, a significant gap persists between technological advancements and clinical integration. We hypothesize that part of this gap arises from the lack of user-friendly interfaces that facilitate the application of these algorithms. Furthermore, few comprehensive solutions exist that provide a fully automated pipeline for SMI determination, including both L3 slice identification and muscle segmentation. Existing tools are often developed within specific cohorts, limiting their applicability for malnutrition screening in broader, unselected patient populations ^6–8^.

This study is a part of a larger research project: the Optimization of the DIAgnosis of SarcoPenia through the automated determination of SMI (ODIASP) study. Our primary objective was to develop and validate an open-access, user-friendly software for the automated determination of the Skeletal Muscle Index (SMI) in clinical research. Additionally, we aimed to evaluate the potential of this tool in identifying the risk of reduced SMI within a cohort of patients managed at a tertiary hospital.

## Methods

### Ethics statement

The ODIASP study was conducted in accordance with the ethical standards laid down in the 1964 Declaration of Helsinki and its later amendments. Ethical approval was obtained on 18 August 2021 by the regional ethics committee (CECIC Rhône-Alpes-Auvergne, Clermont-Ferrand, IRB 5891). In compliance with current French legislation for retrospective studies using clinical data, participants were individually informed that their data could be used for research purposes (in line with the MR-004 CNIL reference methodology). Individuals who objected to the use of their data or those with incomplete or outdated contact information were excluded from the study.

### Study participants

All consecutive participants aged 18 years and older, admitted to Grenoble University Hospital (CHU Grenoble Alpes) between January and December 2018, who underwent at least one CT scan potentially capturing images at the L3 vertebral level and who had a recorded height were retrospectively included in the study. Participants with non-abdominal CT scans, including spinal or thoracic CT scans, were also eligible, as these may contain images at the L3 level. This inclusion was particularly relevant for assessing whether the ODIASP software could effectively detect CT scans that lacked an L3 slice. When multiple CT scans were available for a patient during the inclusion period, the scan most likely to include the L3 slice was selected for analysis. A subset of this population had been previously defined for preliminary research ^9^ and was utilized for validation.

### Data collection

Data were retrospectively collected from the clinical data warehouse (CDW) PREDIMED (French acronym for *Plateforme de Recueil et d’Exploitation des Données bIoMEDicales*), implemented at the Grenoble Alpes University Hospital ^10,11^. All structured data were pseudonymized and CT scans were de-identified. This de-identification of CT scans was carried out by: 1) removing all identifiable Digital Imaging and Communication in Medicine (DICOM) tags following the basic profile recommendations of the DICOM standard^1^, and 2) excluding all derived images (i.e., images where pixel values are derived or computed from other images, such as screenshots or dose reports) based on the ‘ImageType’ DICOM attribute.

The following data were collected

- General data: age, sex, height, weight
- Imaging data: CT scans, including abdominal area, in DICOM format with metadata. Since contrast enhancement does not influence cross-sectional skeletal muscle area, both CT images with and without contrast administration were eligible ^12,13^. CT images could originate from different CT machines (Optima CT660 GE Healthcare, Revolution CT GE Healthcare, Siemens Somatom Definition Edge, Revolution HD GE Healthcare, Revolution EVO GE Healthcare, Toshiba Aquilion and Siemens Somatom Definition AS+).

### Cross sectional muscle area (CSMA) determination

#### Reference Method

All processing was performed on a single GPU machine (NVIDIA TITAN RTX 16 graphics card, a 3.7 GHz CPU, and 64 GB of RAM) with an Intel(R) Xeon(R) W-2135 processor. The third lumbar (L3) vertebra was manually identified by a medical expert on the sagittal reconstruction using the Picture Archiving and Communication System (PACS) in DICOM format. For each participant, a slice approximately halfway along the vertebra was selected for muscle segmentation, with the identification of slices at both the lower and upper extremities. Skeletal muscle was then manually segmented with SliceOmatic Version 5.0 (Tomovision, Canada) according to previously published methods to obtain the CSMA^9^. The abdominal muscles (transversus abdominis, external and internal obliques and rectus abdominis), the paraspinal muscles (erector spinae and quadratus lumborum) and the psoas muscle were segmented using Hounsfield unit (HU) values ranging from -29 to 150 ^14^.

#### ODIASP tool

The ODIASP tool integrates two open-source algorithms: 1) for the automatic selection of a slice at the level of the L3 vertebra, and 2) for the automatic segmentation of skeletal muscle in this slice (Figure 1). For the first step, we employed the L3 slice selection algorithm developed by Bridge et al. ^15^ This algorithm, based on a DenseNet Convolutional Neural Network (CNN), was trained and validated using ODIASP project data through 5-fold cross-validation. A batch size of 64 images, a learning rate of 0.001, and a dropout rate of 0 were selected, while other hyperparameters proposed by Bridge et al. were maintained during the training phase. For the second step, we utilized the AutoMATiCA algorithm developed by Paris et al. ^16^. The proposed U-Net model for automated segmentation of skeletal muscle was previously validated on ODIASP project CT data ^9^.

**Figure 1.**
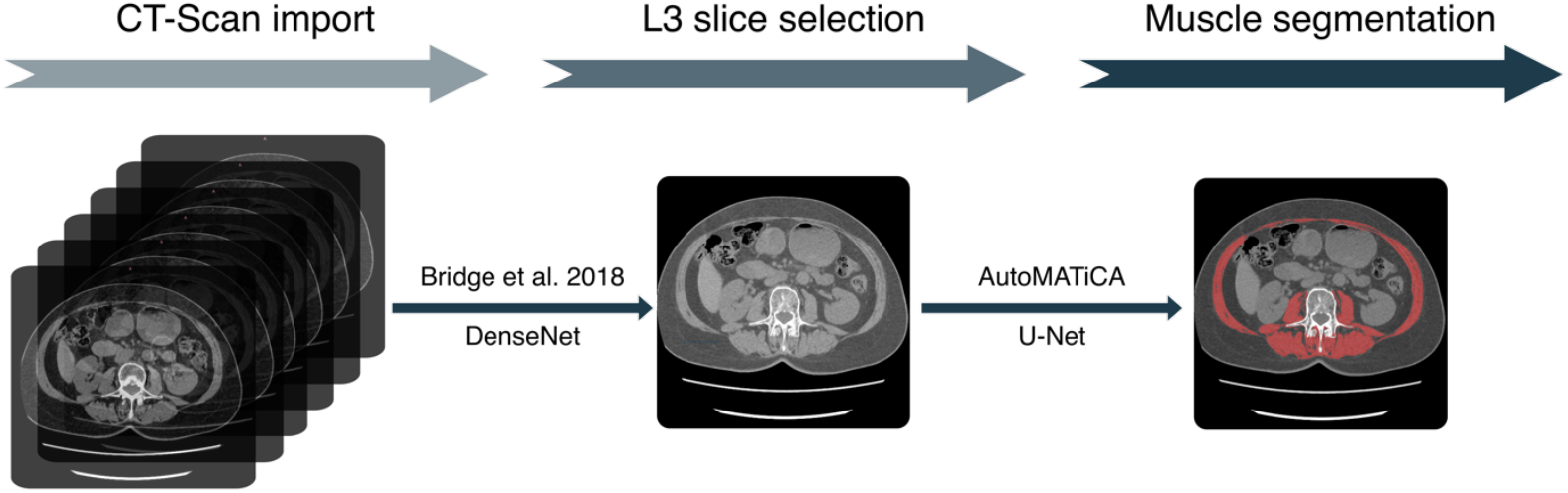
Schematic overview of the architecture of the ODIASP tool

The ODIASP tool is based on two previously published algorithms from Bridge et al. ^15^ and Paris et al. ^16^.

### Statistical analysis

Characteristics of the cohort were presented as medians with interquartile ranges (IQR) or using frequencies and proportions depending on their nature. The agreement between the CSMA values calculated by ODIASP and those obtained using the reference methodology was assessed using the intraclass correlation coefficient (ICC) along with 95% confidence intervals. This analysis was complemented with a mixed regression model. The interpretation of the strength of agreement between the two methods was based on the criteria established by Koo and Li: an ICC value < 0.5 indicated poor reliability, a value between 0.5 and 0.75 indicated moderate reliability, a value between 0.75 and 0.90 indicated substantial reliability, and a value > 0.90 indicated excellent reliability ^17^. For determining the reduction in SMI within our sample, thresholds were derived from the study by van der Werf et al ^18^ and are applicable only to the subpopulation aged 20 to 79 years with a BMI ranging from 17 to 35 kg/m^2^. and were applicable only to the subsample of participants aged 20 to 79 years with a BMI ranging from 17 to 35 kg/m^2^. Statistical analyses were conducted using Stata (version 15) with p-values <0.05 considered statistically significant and without adjustment for test multiplicity.

## Results

### Participant inclusion and characteristics

A total of 3300 participants were included and 2503 participants were analyzed (Figure 2). Their median age was 66 [51–78] years with 1334 (53.3%) males. The median BMI was 24.8 [21.7–28.7] kg/m^2^, with 174 (7.3%) having a BMI < 18.5 kg/m^2^ and 459 (19.3%) classified as obese. The median SMI was 45.1 [38.4–52.5] cm^2^/m^2^. Participants were hospitalized in the following units: day hospital (11.0%), medicine (27.4%), geriatrics (2.8%), surgery (10.4%), intensive care (6.1%), rehabilitation (1.8%), and emergency care (40.5%).

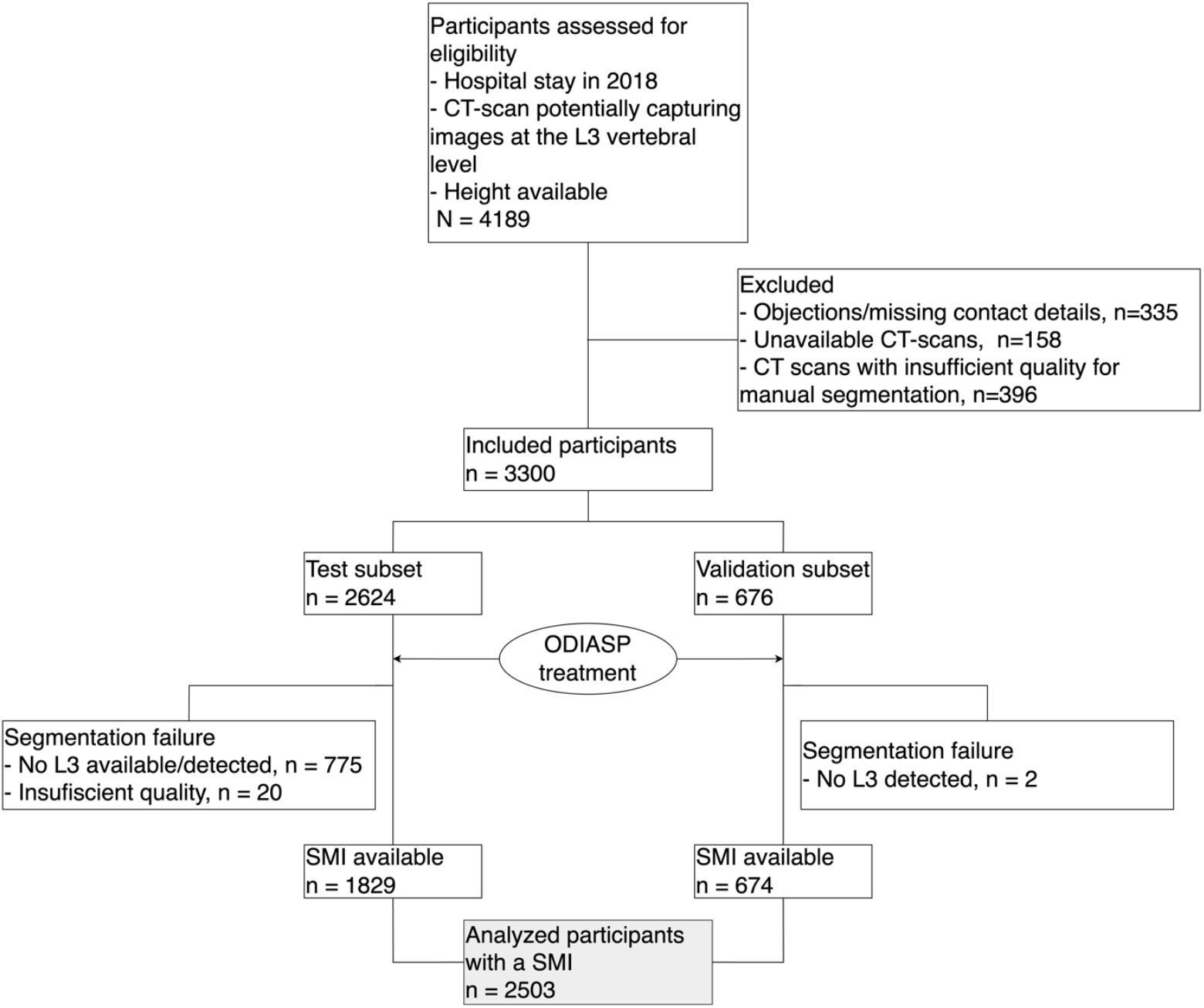

### Validation of the ODIASP tool

The ODIASP tool operates through two main steps: first, identifying a L3 slice and second, segmenting the skeletal muscle (Figure 1). For the first step, we validated the algorithm from Bridge et al. The algorithm correctly identified the L3 slice in 88% of cases (95% CI: 85.49 to 90.50, n=596). For slices outside the L3 level (n=80), the median distance from the chosen L3 slice was -5.6 mm [-17.5 to 12.5], indicating that the slices were, on average, 5.6 mm below the lower extremity of the L3 vertebra. The algorithm for the second step, muscle segmentation, was previously validated externally ^9^.

To validate the entire pipeline, we compared the cross-sectional muscle area (CSMA) obtained using the reference method with that generated by the ODIASP tool in the validation subset. ODIASP failed to provide results for two participants (Figure 2). There was substantial agreement between the reference method and ODIASP (ICC: 0.971; 95% CI: 0.825 to 0.989) (Figure 3). After correcting for systematic errors (a 5.8 cm^2^ [5.4-6.3] overestimation of the CSMA), there was excellent agreement (ICC:0.984, 95% CI: 0.982 to 0.986).

**Figure 3.**
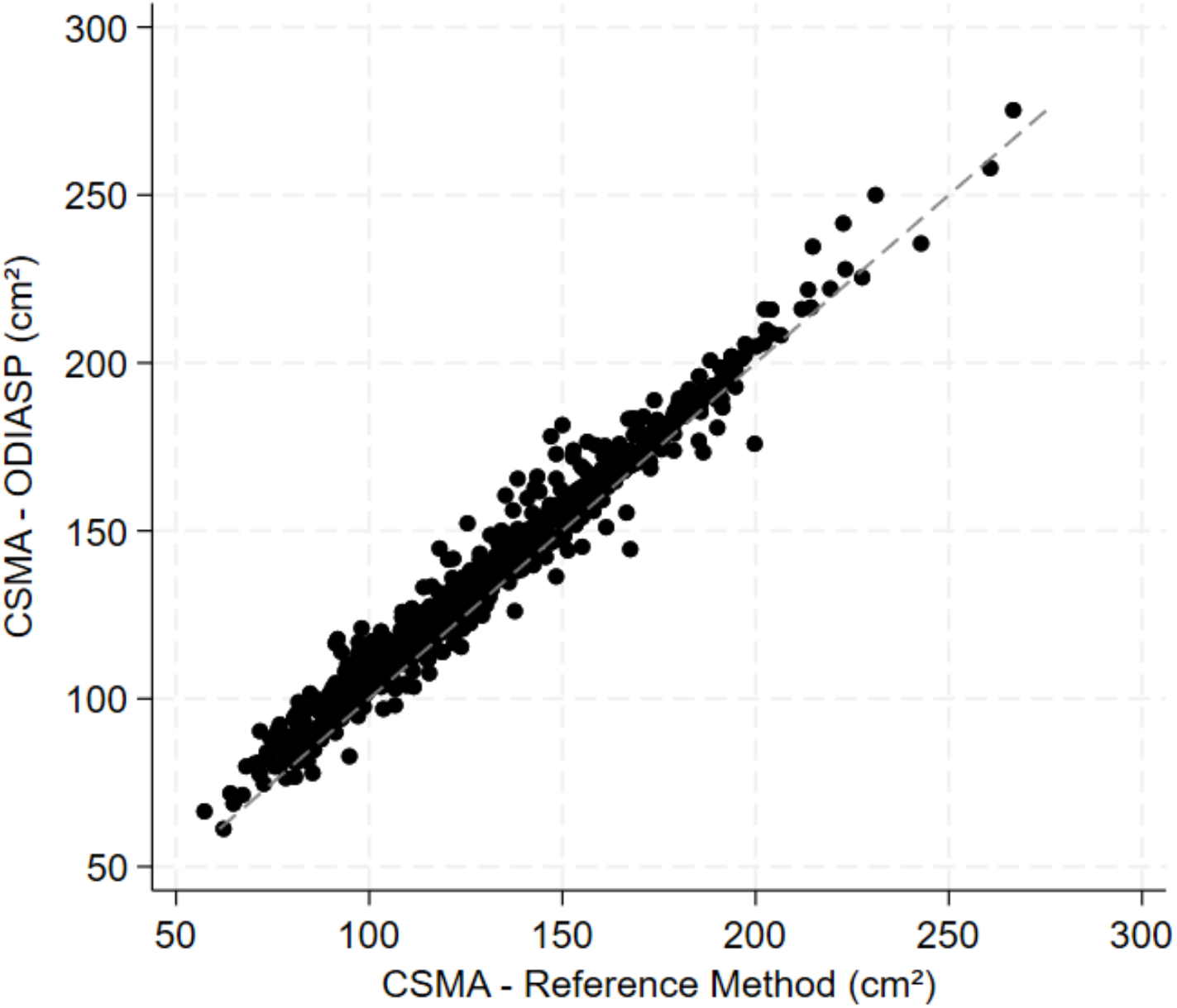
Scatterplot comparing cross-sectional muscle area (CSMA) measurements obtained with the reference method and the ODIASP tool (n = 674).

Each data point represents an individual measurement, and the dashed line signifies perfect agreement between the two methods. The proximity of data points to the dashed line indicates a high degree of agreement.

### An Open-Access and User-Friendly Interface

To facilitate the use of the ODIASP tool, we encapsulated the code into a user-friendly software interface (Figure 4A). The interface displays the sagittal view with the chosen L3 slice localization, the L3 slice itself, and its segmentation, allowing for easy visual validation of results (Figure 4B). This software is freely available at https://odiasp.timc.fr/ (Figure 4C).

**Figure 4.**
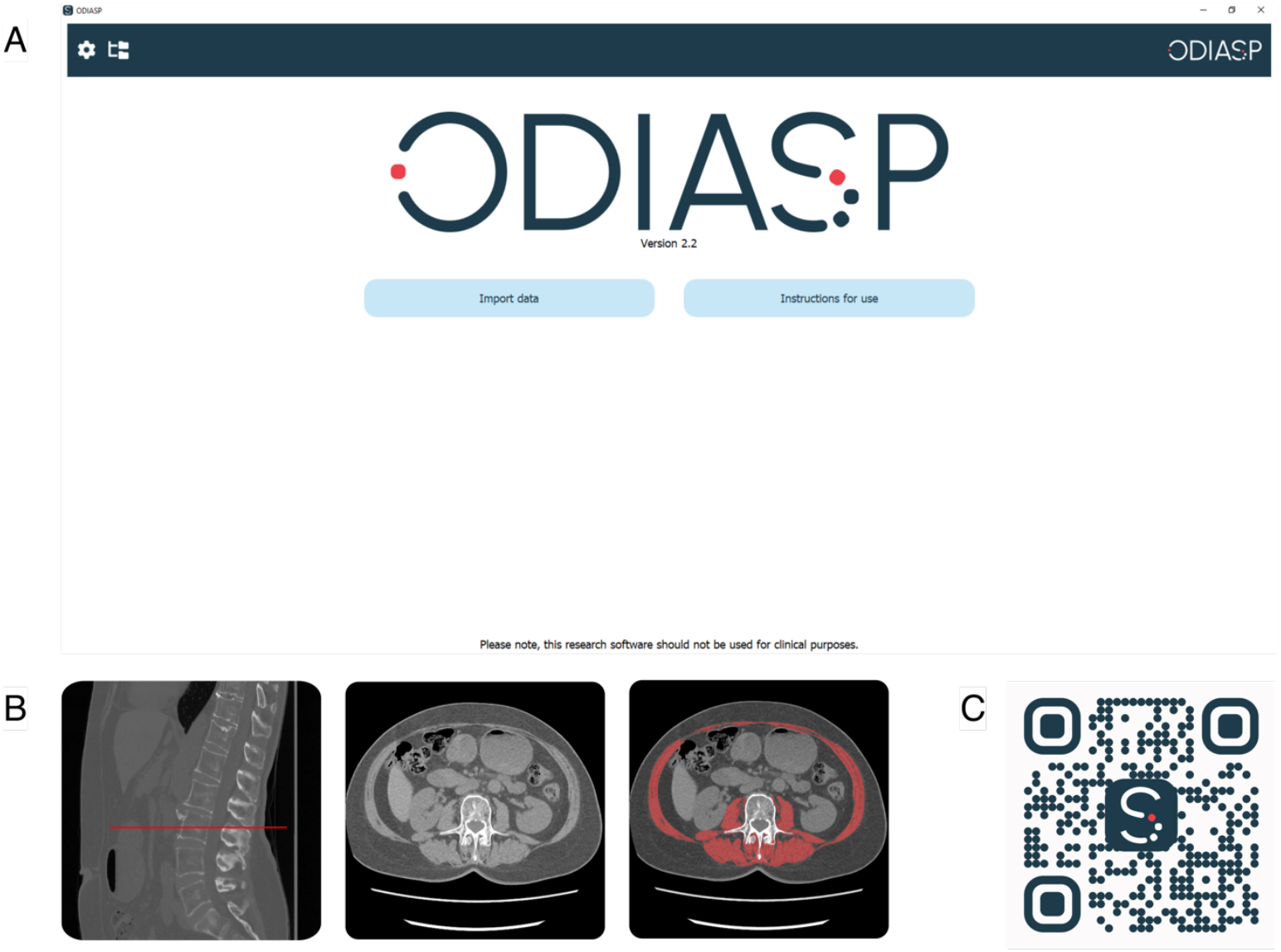
The ODIASP tool: an open access software with user-friendly interface (A) User interface (B) Example of muscle segmentation (C) QR code for downloading the ODIASP Software

ODIASP allows processing of large sets of CT scans without the need for pre-treatment. It takes approximately 4 minutes to analyze one CT scan. In some cases, ODIASP did not correctly identify or segment muscle. However, the interface allows for quick visual inspection of the results if necessary.

### Risk of reduced SMI in a large cohort of hospitalized participants

Based on previously published cut-off values for individuals aged 20–79 years with a BMI between 17 and 35 kg/m^2^, 103 of 937 men (11.0%) and 46 of 699 women (6.6%) were considered to have reduced skeletal muscle mass, representing a total of 9.1% of the participants with an available cut-off ^18^.

## Discussion

This study aimed to develop and validate a tool for the automated determination of SMI at the L3 vertebra level using CT scan in a large population. We integrated AI algorithms into a user-friendly software, making advanced technologies accessible to clinical researchers who may be unfamiliar with them.

### Reliability of the ODIASP automated tool

Most previous studies have focused on tissue segmentation at the L3 level ^16,19,20^, with few addressing the challenge of developing a fully automated pipeline for L3 SMI determination. To our knowledge, we are the first to develop and validate automated AI software for L3 SMI determination in an unselected population of patients managed at a tertiary hospital across various units, including emergency and intensive care. Previous tools have been developed for specific populations, such as cancer patients ^6,7^ or mixed cohorts including cancer, sepsis, and healthy subjects ^8^. Our approach involved utilizing previously published algorithms and validating them in our cohort, demonstrating good reliability across a large sample. We achieved accurate identification of most L3 slices, with 88% correctly positioned at the L3 vertebra, which aligns with previous findings (Delrieu et al.: 91.2% and 74.1% in, two datasets^6^). Slices identified outside of the L3 level showed a median deviation of -5.6 mm [-17.5; 12.5], typically locating them at adjacent vertebrae, given the approximate height of vertebrae (30 mm) and intervertebral disks (10 mm) ^21^. A prior study indicates that CSMA at neighboring vertebrae are relatively comparable ^22^. The reliability of muscle segmentation was previously validated ^9^, and our full pipeline demonstrated substantial to excellent reliability.

### Clinical implications

The prevalence of reduced SMI has mainly been studied in cancer populations ^23–25^. To our knowledge, our study is the first to assess it in a large cohort of unselected patients, with the sole inclusion criteria being the availability of a CT scan. The cut-offs for defining reduced SMI remains controversial, while the cut-offs proposed by Prado et al. ^14^ have been widely used, they are specific to patients with obesity and cancer and can lead to an overestimation of reduced SMI prevalence ^26^. In our study, we referred to the cut-offs by Van der Werf et al.^18^, which are applicable to a Caucasian population and align well with our predominantly Caucasian cohort. Furthermore, Van der Werf et al. provide age- and BMI-specific cut-offs, enhancing the relevance of our findings. Our research indicates that 9% of the patients in our tertiary hospital setting have reduced muscle mass.

### Study limitations

Several limitations should be acknowledged. First, we trained the algorithm of Bridge et al. on our dataset since it was only available in an untrained version. Consequently, our tool lacks full external validation. Second, when testing ODIASP on the analyzed populations, we identified some processing failures. Human oversight remains essential for maintaining data quality; thus, our software provides features for visual manual validation, in line with the recent European Union AI Act emphasizes the necessity of human oversight in algorithmic decision-making ^27^. Third, processing large datasets can be time-consuming; for instance, analyzing 1,000 scans requires just over 24 hours with a standard desktop computer. However, the process is fully automated and runs in the background. Our approach involved testing ODIASP under real-life conditions, using scans with multiple series, including those that did not contain the L3 vertebra, which lengthened the process but reduced the time necessary to preprocess the data. Additionally, it is important to acknowledge that some CT scans may have insufficient quality for accurate segmentation, a factor that should be considered in future clinical research.. Lastly, ODIASP tends to slightly overestimate the CSMA compared to the ground truth. This discrepancy reflects a systematic error comparable to inter-expert variability and could be partly influenced by the overestimation observed with AutoMATiCA or the fact that the ground truth was constructed at our center by only two experts ^9,16^.

### Futures directions

Future work will focus on enhancing the reliability of our software. A strength of this project is our access to diverse medical imaging data through the CDW PREDIMED, allowing for the inclusion of patients from various units and with different pathologies. However, our study is monocentric, which may lead to overfitting and limit generalizability. Traditional training methods can overestimate model accuracy by about 20% due to hidden Data Acquisition Bias-Induced Shortcuts (DABIS) ^28^. Strategies to increase reliability include using external training datasets and applying appropriate data preparation and partitioning techniques (training, validation, and testing sets) to improve generalizability ^29–31^. Additionally, methods to address biases without external datasets can also be applied ^28^. Our objective is to develop extensive cohorts for studying malnutrition and sarcopenia in inpatients. However, many patients do not undergo abdominal CT scans. To address this limitation, we plan to enhance ODIASP’s capabilities to analyze chest CT scans, considering the growing interest in assessing SMI at the T12 vertebra level ^32^. We also plan to validate the segmentation of other tissues— intermuscular, visceral, and subcutaneous adipose tissue—performed in AutoMATiCA before integrating it into ODIASP ^16^. It is important to note that ODIASP is not yet CE marked or FDA approved, limiting its use to clinical research rather than routine practice. Our ultimate goal is to deploy this software in clinical settings after obtaining all mandatory regulatory authorization.

## Conclusion

The results of this study demonstrate that ODIASP is a reliable tool for the automated segmentation of muscles at the L3 vertebra level from CT scans. Integrating validated AI algorithms into a user-friendly software platform can significantly enhance clinical researchers’ ability to assess SMI in large, diverse patient cohorts. We anticipate that our research will facilitate a deeper understanding of the impact of reduced SMI, ultimately contributing to improve patient outcomes through more accurate malnutrition and sarcopenia assessments.

## Data Availability

Data produced in the present study are available upon reasonable request to the authors (with the exception of the CT scan data, which may be considered identifying)

## Funding

This work was supported by a grant from the Regional Delegation for Clinical Research of the University Hospital Grenoble Alpes in 2019 and MIAI@Grenoble Alpes, (ANR-19-P3IA-0003). The funding bodies did not have any involvement in the design/conduct of the research, in data analysis/interpretation, or in writing/approval of the manuscript.

## Conflict of interest Statement

Katia Charrière, Antoine Ragusa, Béatrice Genoux, Antoine Vilotitch, Svetlana Artemova, Charlène Dumont, Paul-Antoine Beaudoin, Pierre-Ephren Madiot, Gilbert R. Ferretti, Ivan Bricault, Jean-Luc Bosson, Eric Fontaine, Alexandre Moreau-Gaudry, Joris Giai and Cécile Bétryhave no relevant conflict of interest to disclose.

## Ethical guidelines statement

The ODIASP study was conducted in accordance with the ethical standards laid down in the 1964 Declaration of Helsinki and its later amendments. Ethical approval was obtained on 18 August 2021 by the regional ethics committee (CECIC Rhône-Alpes-Auvergne, Clermont-Ferrand, IRB 5891). In compliance with current French legislation for retrospective studies using clinical data, participants were individually informed that their data could be used for research purposes (in line with the MR-004 CNIL reference methodology)

## Footnotes

1 https://dicom.nema.org/medical/dicom/current/output/html/part15.html#sect_E.1.1

## Notes

### Competing Interest Statement

The authors have declared no competing interest.

### Author Declarations

Ethical approval was obtained on 18 August 2021 by the regional ethics committee (CECIC Rhone-Alpes-Auvergne, Clermont-Ferrand, IRB 5891)

